# Abortion laws and severity of abortion-related morbidities: A cross-sectional analysis of the WHO multi-country survey on abortion (WHO MCS-A) in sub-Saharan Africa

**DOI:** 10.1101/2025.09.25.25336692

**Authors:** Blake Erhardt-Ohren, Hedieh Mehrtash, Adama Baguiya, Kwame Adu-Bonsaffoh, Philip Govule, Ausbert Thoko Msusa, Folasade Adenike Bello, Zahida Qureshi, Rachidatou Compaore, Caron Rahn Kim, Özge Tuncalp, Ndola Prata

**Affiliations:** Bixby Center for Population, Health and Sustainability, School of Public Health, University of California, Berkeley, Berkeley, USA; UNDP/UNFPA/UNICEF/WHO/World Bank Special Programme of Research, Development and Research Training in Human Reproduction (HRP), Department of Sexual and Reproductive Health and Research, WHO, Geneva, Switzerland; Unité de recherche de Kaya, Institut de Recherche en Sciences de la Santé (IRSS), Ouagadougou, Burkina Faso; Department of Obstetrics and Gynecology, University of Ghana Medical School/Korle-Bu Teaching Hospital, Accra, Ghana; Department of Epidemiology and Disease Control, School of Public Health, University of Ghana, Accra, Ghana; Centre for Reproductive Health, College of Medicine, University of Malawi, Blantyre, Malawi; Department of Obstetrics and Gynaecology, University of Ibadan, Ibadan, Nigeria; Department of Obstetrics and Gynaecology, University of Nairobi, Nairobi, Kenya

**Keywords:** abortion, abortion complications, post-abortion care, abortion legal framework, abortion policy, reproductive rights, sub-Saharan Africa

## Abstract

Less and least safe abortion rates are higher in countries with restrictive abortion laws than in countries with less restrictive laws. Women seek out and procure services regardless of the legal status of abortion – in countries with restrictions, this means they may seek abortions outside of the legal system, which are more likely to be unsafe. In this study, we investigate whether the severity of abortion-related severe morbidity differs with varying levels of abortion legal restrictions.

This is a secondary analysis using data from the WHO’s MCS-A study, which was a cross-sectional study of 11 countries in sub-Saharan Africa. Abortion-related morbidity was categorized by severity. We divided abortion laws into three categories: abortion on request, abortion for health indications, and abortion prohibited altogether. We constructed a logistic regression model to understand the effect of restrictive abortion laws on the odds of more severe complications compared to less severe complications. We included 7,475 women across 210 health facilities. More women lived in countries with abortion for health indications (74%) than countries with abortion on request (19%) or countries where abortion was prohibited altogether (8%). Most women (91%) had less severe abortion-related complications. Women living in countries where abortion is legal for health indications were significantly more likely to experience severe complications than those living where abortion is available on request (1.10 OR, 95% CI 1.07-1.12). Women in countries where abortion is prohibited altogether similarly had significantly higher odds of more severe abortion complications than those living in countries where abortion is available on request (1.10 OR, 95% CI 1.07-1.14). In sub-Saharan Africa, abortion complications are statistically significantly higher in countries with restrictions compared to those with abortion on request. While easing restrictions on abortion could significantly reduce abortion complications, initiatives to increase access to timely and quality comprehensive abortion care is an urgent matter, as it would likely improve outcomes. These initiatives should focus on raising awareness about the law, values clarification for health workers and policy makers, and targeting other barriers at the facility– and community-level that might delay or prevent care.

## Background

In sub-Saharan Africa, 42% of pregnancies were unintended from 2015-2019 and 37% of these pregnancies ended in abortion [1]. Induced abortion rates are highest in countries that restrict abortion access and lowest in countries that guarantee access to reproductive health services, including where induced abortion is broadly available [2]. The World Health Organization (WHO) defines “least safe” abortions as those that are not provided by trained health care workers and not performed using a WHO-recommended procedure appropriate to pregnancy duration, whereas “less safe” abortions fall into one of those categories [3]. Globally, less and least safe abortion rates are higher in countries with restrictive laws than in countries with less restrictive laws [4]: 77% of the abortions in sub-Saharan Africa in 2015-2019 were estimated to be unsafe [1]. Women seek out and procure services regardless of the legal status of abortion – in countries with restrictions, this means they may seek abortions outside of the legal system, which are more likely to be unsafe [2].

Abortion laws vary greatly across the 54 countries of Africa. In 2024, eight countries allow abortion on request, three countries permit abortion on socioeconomic grounds, 25 to preserve health, and twelve grant abortion to save the pregnant woman’s life [5]. In only six countries in Africa is abortion prohibited altogether. An additional 24 countries, excluding those with abortion on request, allow abortion in cases of rape. In Africa in 2023, 100% of countries had criminal penalties for abortion health workers, 98% of countries had criminal penalties for abortion “assistors”, and 91% for abortion seekers, for abortions sought, procured, or provided outside of the legal framework of the country [6].

Despite abortion being legal for at least one indication in most countries in Africa, not all pregnant women and girls are aware of the service [7, 8], and health workers may limit who is able to access services based on their knowledge of the law or personal beliefs [9, 10]. Other barriers, such as cost of services, may prevent access to abortion [11]. The criminalization of abortion services and their lack of accessibility negatively impact health outcomes and health systems [12].

Previous studies have explored the relationship between the availability of abortion services and abortion safety (least safe, less safe, safe) across countries [13–15]. A small amount of ecological research has linked abortion law and overall abortion-related morbidity and mortality, both of which found that decriminalization may save lives [16–17]. The purpose of this study is to investigate whether the severity of abortion-related morbidities differs across countries with varying levels of abortion restrictions (abortion on request, abortion for health indications, abortion altogether prohibited), using the data from the WHO Multi-country Survey on Abortion (WHO MCS-A).

## Methods

### Study design

This is a secondary analysis of the WHO’s Multi-Country Survey on Abortion (WHO MCS-A) [18]. Briefly, the MCS-A survey utilized prospective data collection, with multi-stage sampling at country, province, and all facility levels covering Africa, Latin America, and the Caribbean region. This analysis includes eleven countries in Africa (Benin, Burkina Faso, Chad, Democratic Republic of the Congo, Ghana, Kenya, Malawi, Mozambique, Niger, Nigeria, Uganda).

### Data source

Health facilities were eligible for inclusion in the study if they had more than 1,000 deliveries per year, to provide a robust sample size, had signal functions for emergency obstetric care, i.e., implemented specific activities to prevent maternal and neonatal morbidity and mortality, provided abortion and/or post-abortion care to the extent of the law, and had, on average, at least ten post-abortion clients per month. Data collection comprised three months in each country between February 2017 and April 2018.

All women presenting to each facility with signs and symptoms of abortion complications or miscarriage, or death at facility discharge from abortion-related complications or miscarriage were included in the study. Research assistants abstracted information from medical records for each eligible woman. The full study protocol and main results findings are published elsewhere [18–21].

### Definitions and categories

We categorized women into two groups based on the severity of their abortion complications: mild and moderate complications, which we call *“less severe complications”* and potentially life-threatening complications, near-misses, and deaths *which we call “more severe complications”* [18–21]. Included within mild complications were vaginal bleeding, open cervix, abnormal vital signs, uterine tenderness, abnormal mental state, abnormal abdominal, abnormal appearance, cervical motion tenderness, foul smelling vaginal discharge, evidence of foreign body, and adnexal mass. Moderate complications included bleeding symptoms, suspected intra-abdominal injury, and infection. Near-miss complications refer to cardiovascular, respiratory, renal, coagulation, neurologic, hepatic, or uterine organ dysfunction. Potentially life-threatening complications encompass severe hemorrhage, severe systemic infection, and uterine perforation.

We classified each country’s abortion laws according to the categories used by the Center for Reproductive Rights: i) on request; ii) broad social or economic grounds iii) to preserve health; iv) to save a person’s life; and v) prohibited altogether [5]. In this analysis, we combined abortion to preserve health and abortion to save a person’s life into one category: “abortion for health indications”.

We explored covariates at country level, such as: whether misoprostol is approved for post-abortion care treatment in the country (yes/no), facility location (urban, peri-urban or rural), and at individual level: age (<25 years, 25+ years), marital status (married, single, separated/divorced/widowed), level of education (none, primary completed, secondary completed), gestational age (first trimester [<13 weeks], second trimester [13-27 weeks]), previous births (0, 1+), and previous induced abortions and miscarriages (0, 1+). These variables were selected a priori based on theoretical associations between them and the outcome of interest. We removed an occupation variable due to collinearity with the level of education variable.

### Statistical analysis

We used R statistical package version 4.1.2 for statistical analyses [21]. We removed records with missing information for any variables of interest. We conducted bivariate analyses to assess differences between women with less severe and more severe complications according to country abortion law restrictiveness. We then constructed logistic regression models (unadjusted and adjusted) to assess the likelihood of more severe complications given abortion law restrictiveness. Adjusted models included variables that were associated with the outcome at p<0.20 in the bivariate analysis. We utilized log-likelihood test ratios to compare the fit of the full model against reduced models, wherein each predictor variable was removed to assess fit. In our final model, we included only the variables that contributed to the explanatory power of the model. We established statistical significance of variables in the final model at p<0.05. For all logistic regression models, we present unadjusted odds ratios, adjusted odds ratios, and their 95% confidence intervals.

### Ethical considerations

This study was approved by the World Health Organization Ethical Review Committee (#0002699). We also received ethics approval in each country: Benin (Le Comité National d’Ethique pour la Recherche en Santé), Burkina Faso (Le Ministère de la Recherché Scientifique et de l’Innovation), Chad (Ministère de l’Enseignement Supérieur et de la Recherche Scientifique), the Democratic Republic of Congo (Ecole de Santé Publique Comité d’Ethique), Ghana (Ethical Review Committee of the Ghana Health Service and Ethical and Protocol Review Committee of the College of Health Sciences, University of Ghana), Kenya (Kenyatta National Hospital/University of Nairobi (KNH/UoN) Ethics and Research Committee), Malawi (College of Medicine Research Ethics Committee (COMREC)), Mozambique (Comité Nacional de Bioetica para e Saude, Ministerio de Saude), Nigeria (Federal Capital Territory Health Research Ethics Committee; Research Ethical Review Committee, Oyo State and State Health Research Ethics Committee of Ondo State) and Uganda (Mulago Hospital Research Committee; Uganda National Council for Science and Technology). All women participating in exit interviews provided informed consent.

## Results

We used data from 210 health facilities across 11 countries in Africa. At the time of data collection, one country provided abortion on request (Mozambique), nine countries provided abortion for health indications (Benin, Burkina Faso, Chad, Ghana, Kenya, Malawi, Niger, Nigeria, and Uganda), and the Democratic Republic of Congo prohibited abortion altogether. No countries in our analysis allowed abortion on broad socioeconomic grounds. Only three countries did not have misoprostol approved for use (Burkina Faso, Niger, and Chad). A map detailing the countries included in the analysis is available in *Fig 1*.

**Fig 1.**
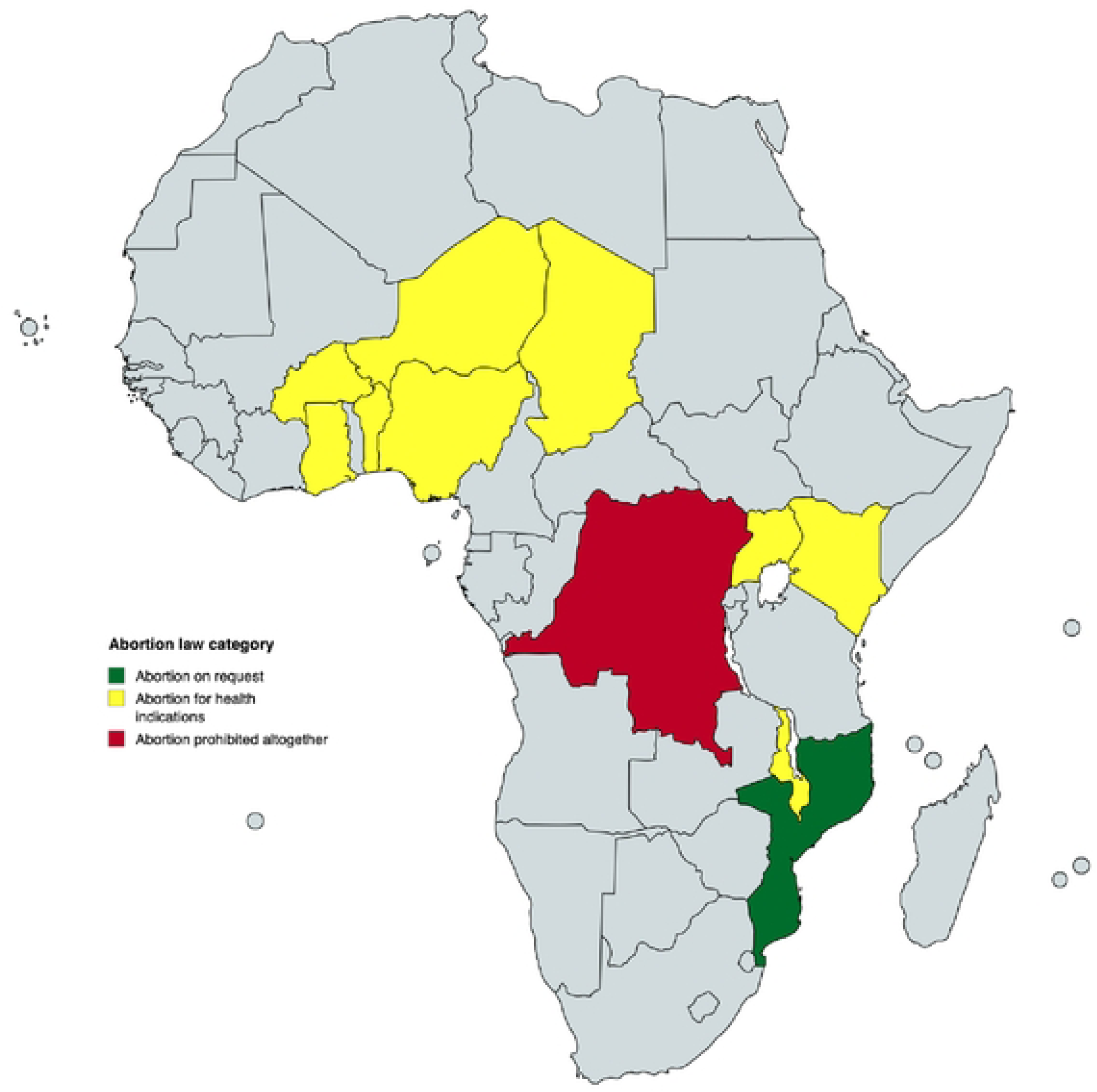
Abortion law categories of eleven countries included in the analysis, at the time of data collection, 2017-2018 [22].

### Study participants

The study collected data from 13,657 women with abortion-related complications, but our final sample includes 7,475 women (55%) with complete records for variables of interest (see *Fig 2* for a flow chart of all participants; see *S1 Table* for a table containing results from a bivariate analysis comparing women with and without complete records for the variables of interest). Approximately four-fifths (79%) of the women resided in countries with abortion for health indications, while approximately one eighth lived in countries with abortion on request (13%), and 8% in the Democratic Republic of Congo, where abortion is prohibited altogether. 20% of women lived in countries where misoprostol is not approved for post-abortion care. The majority of women sought care in urban locations (72%). About one quarter of the women (26%) were less than 25 years old, 82% were married or cohabiting, and they were evenly divided across educational levels (no education: 35%, primary completed: 32%, secondary completed: 33%). The majority (65%) of women had a previous birth (91%) and 38% had a previous abortion. Regarding the terminated pregnancy, 61% of women were in their first trimester. Most women (90.6%) had less severe abortion-related complications; there were no deaths in our final sample of women. The following covariates were associated with abortion complication severity in bivariate analyses at p<0.20: abortion law category, misoprostol approved for use, facility location, age, marital status, education level, and gestational age. Detailed characteristics of the study sample are available in *Table 1*.

**Fig 2.**
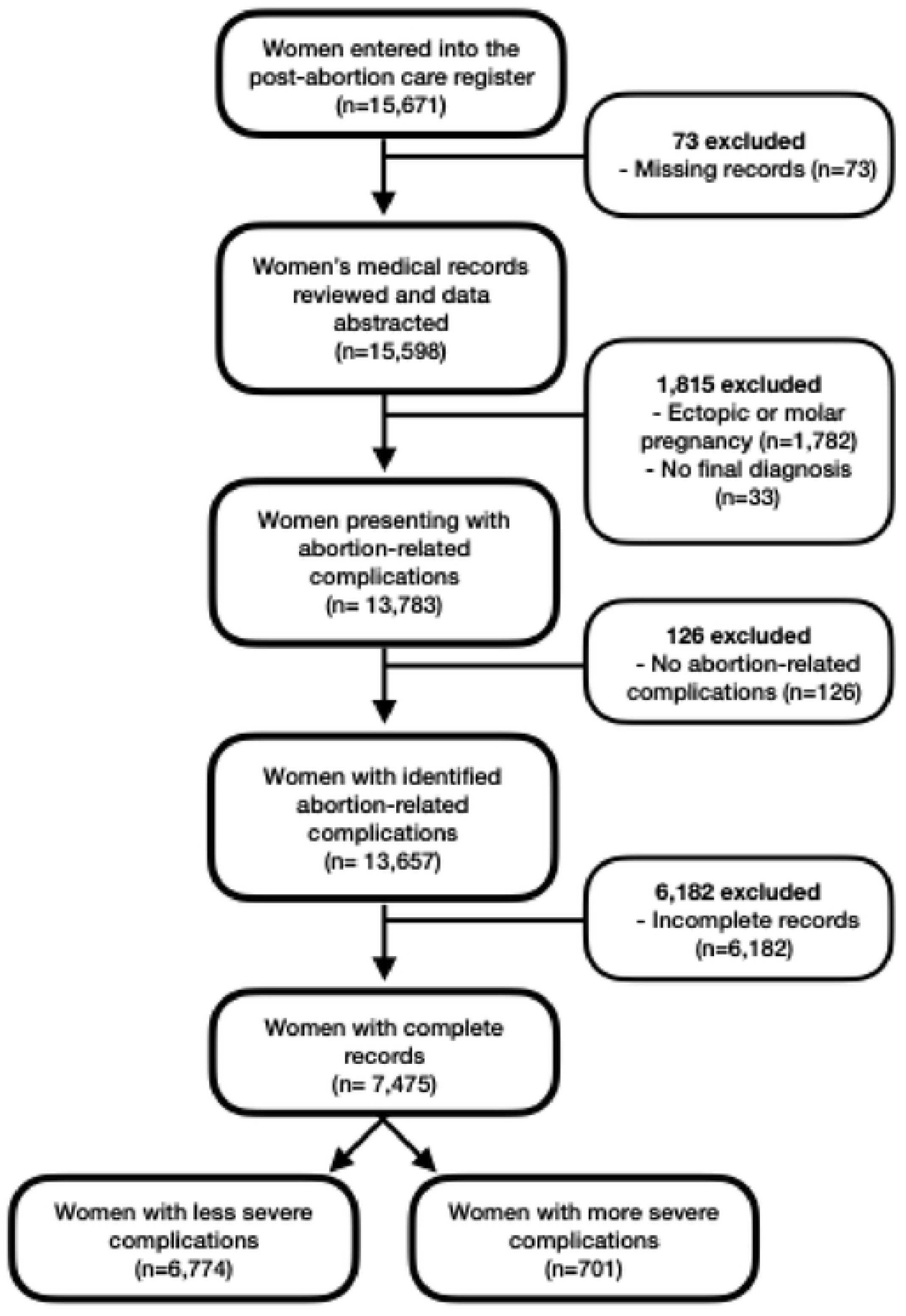
Flow chart of participants in the study sample.

**Table 1.**
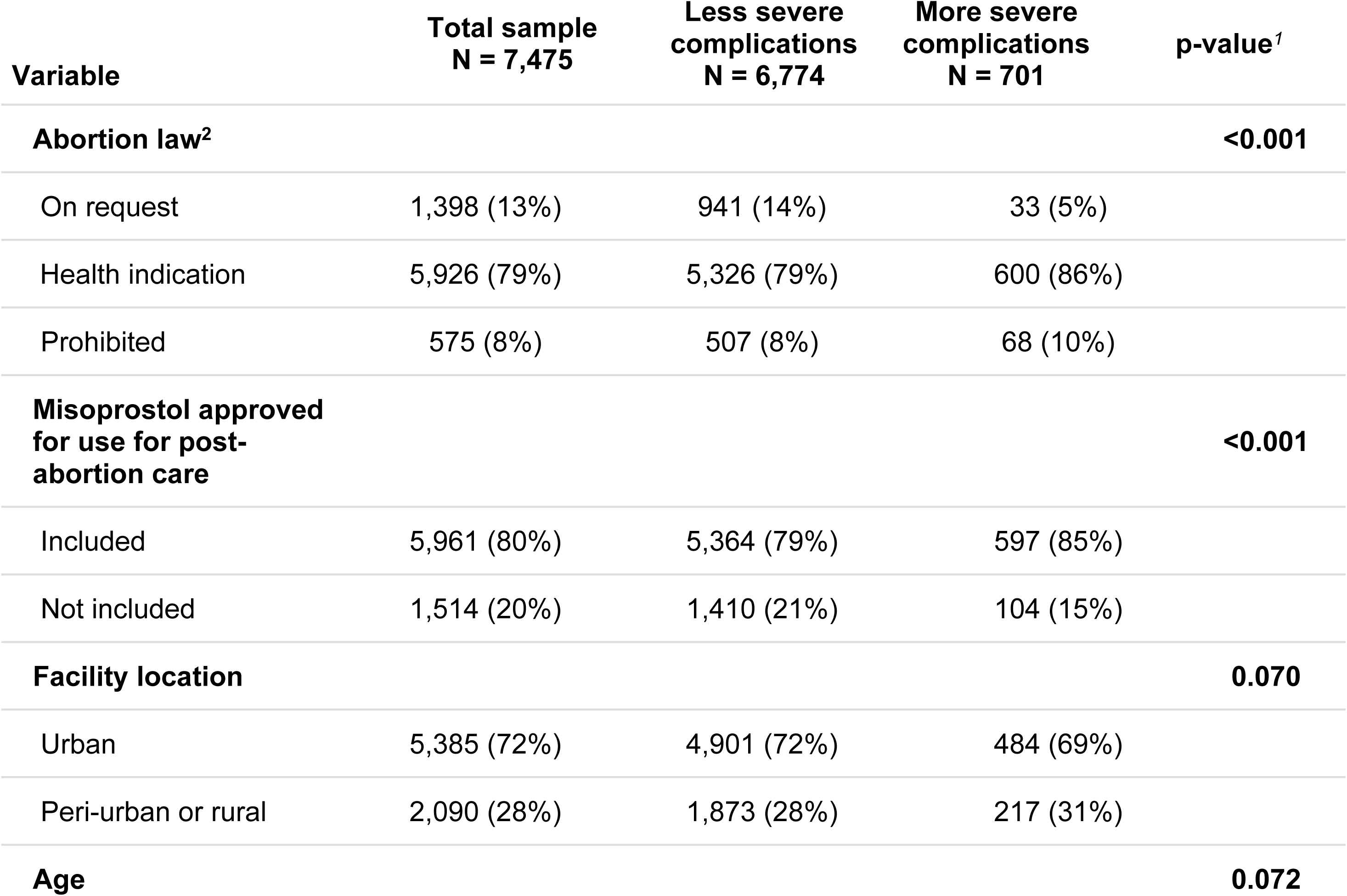
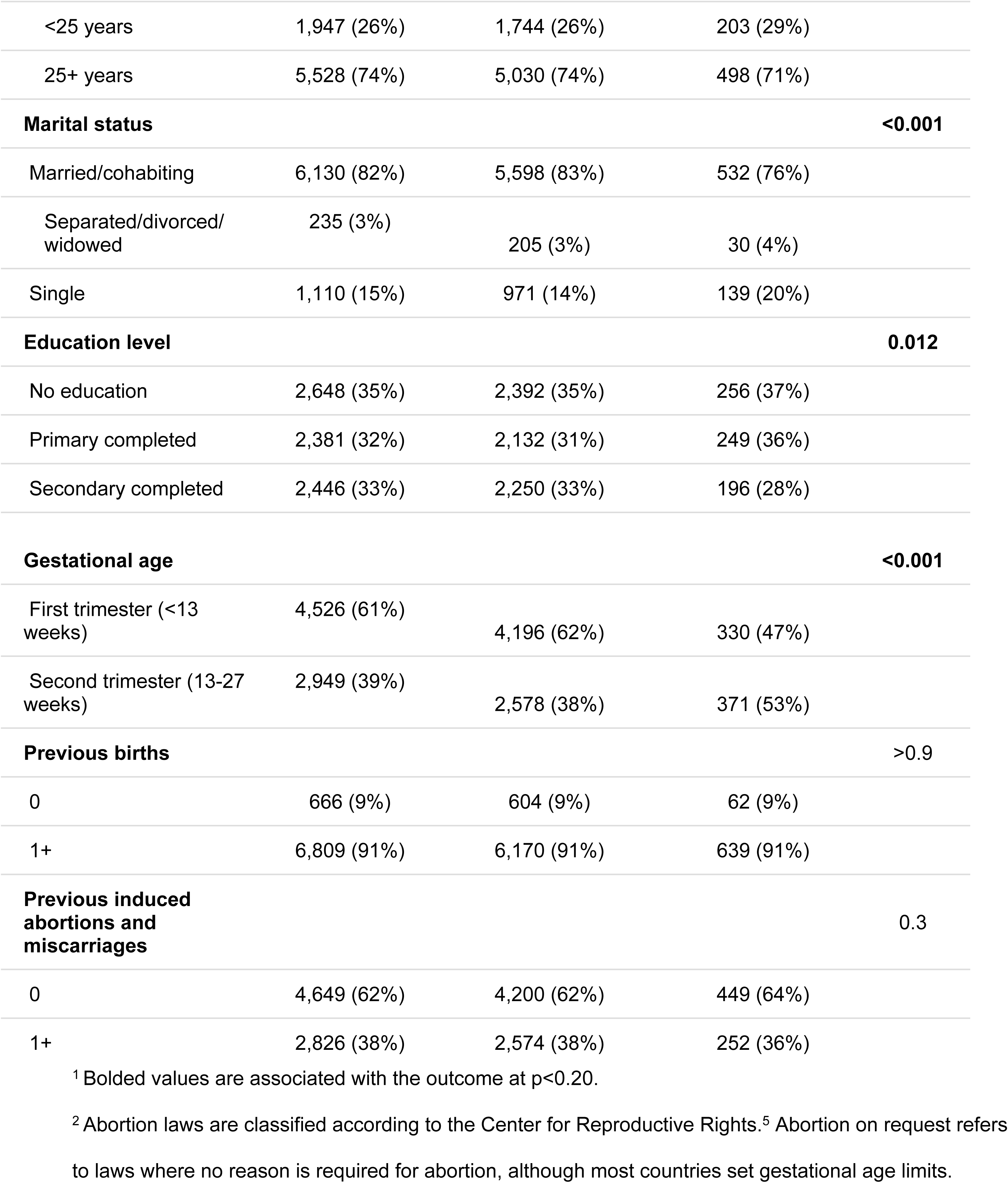
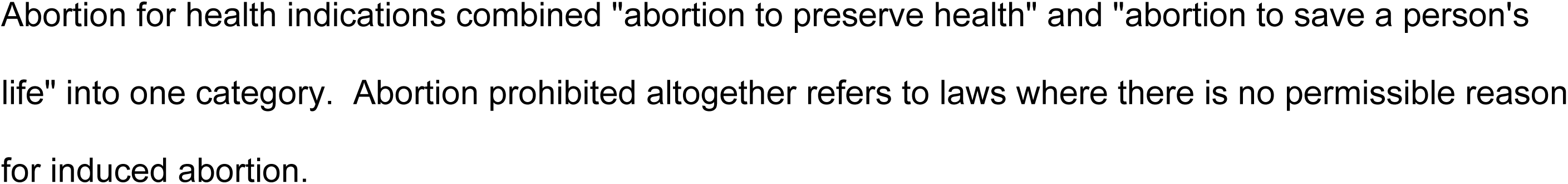
Characteristics of women according to abortion complication severity.

In the unadjusted logistic regression models, women in countries with more restrictive abortion laws had significantly greater odds of more severe abortion complications than in countries where abortion is available on request: unadjusted OR 1.07 (95% CI 1.05-1.09) in countries with abortion allowed for health indications and unadjusted OR 1.09 (95% CI 1.06-1.12) in countries with abortion prohibited altogether.

To select the final model, we initially included all variables of interest (abortion law category, misoprostol approved for use, facility location, age, marital status, education level, and gestational age) that were significantly associated with the outcome of interest at p<0.20. After running log-likelihood ratio tests, we removed the following variables that did not contribute significantly at p<0.05 to the model’s explanatory power: age and facility location. The adjusted model’s results were also statistically significant with a slight increase in effect size for both categories of abortion restrictions compared to abortion on request. The odds of more severe abortion complications in countries with abortion for health indications was 1.10 times (95% CI 1.07-1.12) that of those in countries where abortion is available on request and women in countries with abortion prohibited altogether had 1.10 (95% CI 1.07-1.14) times higher odds of more severe abortion complications than women in countries where abortion is available on request. The results of the unadjusted and adjusted models are in *Table 2*.

**Table 2.** Logistic regression results of the association between restrictive abortion laws and more severe versus less severe abortion-related complications.

| Predictor | Crude odds ratio<br>(confidence interval) | P-value<sup>2</sup> | Adjusted odds ratio<br>(confidence interval)<sup>1</sup> | P-value<sup>2</sup> |
| --- | --- | --- | --- | --- |
| Abortion on request | — | — | — | — |
| Abortion for health indications | 1.07 (1.05-1.09) | **<0.001** | 1.10 (1.07-1.12) | **<0.001** |
| Abortion prohibited altogether | 1.09 (1.06-1.12) | **<0.001** | 1.10 (1.07-1.14) | **<0.001** |

<sup>1</sup> We included the following variables in the adjusted logistic regression model: marital status (married, single, separated/divorced/widowed), level of education (none, primary completed, secondary completed), gestational age (first trimester, second trimester), previous births (0, 1+), and previous induced abortions and miscarriages (0, 1+).
<sup>2</sup> Bolded values are significant at p<0.05.

## Discussion

In this manuscript, we explore the relationship between abortion laws (abortion on request, abortion for health indications, and abortion prohibited altogether) and abortion-related morbidity across 210 facilities in 11 countries in 2017-2018. Across countries in our study, we observed that abortion legal restrictions are significantly associated with the severity of abortion complications. We found that there are statistically significantly higher odds of more severe abortion-related complications than less severe complications in countries with restrictive abortion laws. Although, in our analysis we have two countries whose abortion legal frameworks have changed since data collection (Benin [23] and DRC [24]), our results are still valid as an indication of how abortion laws are related to the severity of abortion-related complications.

In the context of sub-Saharan Africa in 2019, 92% of women of reproductive age, 15-49 years, lived in countries with moderately or highly restrictive laws [1]. And while maternal mortality around the world decreased by 34% from 2000 to 2020, progress was slower in sub-Saharan Africa, which accounted for 70% of all global maternal deaths in 2020 [25]. In 2010-2014, a Guttmacher Institute-WHO research collaboration estimated that 77% of abortions in sub-Saharan Africa were unsafe, compared to 45% globally [2]. Abortion laws vary dramatically globally [26], and previous research in Nigeria, Cote d’Ivoire, and India confirms that abortion safety varies by abortion legal context: significantly more abortions were less or least safe in the countries with abortion to save a woman’s life than in the country with abortion available on board social or economic grounds [13]. These less safe abortions are more likely to result in abortion complications [27]. Ultimately, this contributes directly to maternal death: in 2019, sub-Saharan Africa had the highest abortion case-fatality rate of any world region, contributing 15,000 deaths each year [1]. Without intervention, the issue of preventable abortion-related morbidity and mortality will continue, impacting women around the world’s right to health and life [28].

Between 2008 and 2023, the population of sub-Saharan Africa grew by approximately 50% and it is projected to increase until at least 2040 [29]. While the population grows, the abortion rate is likely to remain the same as it has for the past quarter decade — around 33 abortions each year per 1,000 women aged 15-49 years [29]. This means the total number of abortions, and therefore the total number of unsafe abortions, and related morbidity and mortality is likely to continue to increase in the absence of any legal changes.

There are many potential solutions to address this issue: the most likely to cause large-scale decreases in abortion-related complications is to liberalize abortion laws across sub-Saharan Africa. While several African countries changed their abortion laws in response to the Maputo Protocol, most remain highly restrictive. Building on the evidence that restrictive laws present barriers to access, the WHO Abortion care guideline includes seven law and policy recommendations covering the issues of criminalization, grounds-based approaches, conscientious objection, third party authorization, gestational age limits, provider restrictions, and mandatory waiting periods.^3^ Amending laws and policies is an iterative process, but active steps may be taken to liberalize abortion laws throughout the process [30]. Decriminalizing induced abortion, changing the application of the law to improve access to high quality services, and developing national guidelines for comprehensive abortion care that align with WHO’s Abortion care guideline are highly effective ways to decrease morbidity and mortality [3], and is possible through political coalitions, as demonstrated by Ethiopia in 2005 [31] and Rwanda in 2012 [32]. Changes in application of the law may also improve abortion outcomes.

While legal reform may decrease the number of abortion complications, it doesn’t necessarily guarantee access to quality abortion care: comprehensive policy approaches must address healthcare system capacity, health worker training and support, financial accessibility, geographic distribution of services, public education and awareness, and stigma reduction [3]. A 2023 study in Pakistan, where abortion is available to preserve health, demonstrated that clinical training, community engagement, and engagement of PAC recipients in integrated maternal health and family planning could decrease unintended pregnancies, unsafe abortion, and maternal mortality, and was a model that could be feasibly implemented in health systems [33]. Induced abortion on request is a long-term solution, but there are opportunities for changes in the law or its application to improve safe abortion access, and for continuous improvements in PAC provision to ensure timeliness and quality of care to save lives and prevent harm from abortion complications.

## Strengths and limitations

To our knowledge, this is one of the few studies to look at the association between abortion laws, across several countries with different law severity, and abortion morbidity using individual-level data. We use national-, facility-, and patient-level data to parse out the association between more restrictive abortion laws and the likelihood of more severe abortion outcomes with a large cohort of over 7,000 women across 210 facilities and 11 countries. However, there are limitations that affect interpretation of study results. We are unable to adjust for the degree to which the laws are implemented, or not implemented, in each facility in which we collected data, and the study population does not include a country with abortion available on broad socioeconomic grounds. While we aim to understand the relationship between abortion laws and abortion complications, our analysis does not include women who received abortions and did not experience complications. Ideally, we would have been able to include all women who did and did not have abortion-related complications. We also recognize that individuals with severe abortion complications are more likely to attend higher level health facilities, such as the higher-volume facilities in this study. However, we do not believe that if we had included lower-level facilities, the results would be significantly difference because patients with severe abortion complications are referred to higher-level facilities.

Additionally, the project collected data from 13,657 women, but we included only 7,475 complete records (55%) and most variables significantly differed between women with complete and incomplete records (see *S1-S2 Tables*). This is not unusual in low– and middle-income countries, where inadequate training, limited infrastructure, and organizational constraints prevent consistent medical record-keeping, especially on sensitive subjects, such as abortion [34]. A sensitivity analysis revealed no significant differences in results using the complete or incomplete and complete records. Despite these limitations, this study clearly indicates an association between the legal framework for abortion and the severity of abortion-related complications.

## Conclusions

This study demonstrates that in sub-Saharan Africa, restrictive abortion laws are associated with increased odds of potentially life-threatening complications and near-misses among women seeking PAC. Initiatives to increase accessible legal abortion service provision in countries with indications for abortion are likely to improve these outcomes. These initiatives should focus on education about the law, values clarification for health workers and policy makers, and targeting other barriers at the facility– and community-level. Policy initiatives to expand access to services may reduce excess abortion-related complications. Liberalizing abortion laws would increase women’s abilities to exercise their human rights and lead to reductions in preventable complications.

## Data Availability

The data that support the findings of this study are available from the UNDP/UNFPA/UNICEF/WHO/World Bank Special Programme of Research, Development and Research Training in Human Reproduction (HRP), Department of Sexual and Reproductive Health and Research team at the World Health Organization but restrictions apply to the availability of these data, which were used under license for the current study, and so are not publicly available. Data are available from the authors upon reasonable request and with permission of the WHO.

## Acknowledgements

We are grateful to the women who participated in the study. In addition, we would like to thank the other members of the WHO MCS-A team who conceived of, designed, and implemented the project. We would also like to thank the participants of the IUSSP International Seminar on *Improving measurement of abortion incidence and safety: Innovations in methodology and recent empirical studies* for their comments and questions that strengthened this manuscript.

## List of abbreviations

● MCS-A: Multi-Country Study on Abortion
● WHO: World Health Organization

## Consent for publication

Not applicable

## Competing interests

The authors declare that they have no competing interests.

## Funding

This particular study was not funded, but the parent study was funded by UNDP/UNFPA/UNICEF/WHO/World Bank Special Programme of Research, Development and Research Training in Human Reproduction (HRP); Department of Sexual and Reproductive Health and Research; World Health Organization.

## Authors’ contributions

HM, ÖT, AB, KAB, PG, ATM,FAB, ZQ, RC, and CRK all contributed to the design of the WHO MCS-A project and data collection for the project. BEO and NP conceptualized, designed, and implemented this secondary analysis with input from the other co-authors who assisted with interpretation. BEO and NP drafted this manuscript. All authors read and consented to this manuscript’s submission for publication. Disclaimer: The authors alone are responsible for the views expressed in this article and they do not necessarily represent the views, decisions or policies of the institutions with which they are affiliated.

## Notes

### Competing Interest Statement

The authors have declared no competing interest.

### Author Declarations

This study was approved by the World Health Organization Ethical Review Committee (#0002699). We also received ethics approval in each country: Benin (Le Comité National d’Ethique pour la Recherche en Santé), Burkina Faso (Le Ministère de la Recherché Scientifique et de l’Innovation), Chad (Ministère de l’Enseignement Supérieur et de la Recherche Scientifique), the Democratic Republic of Congo (Ecole de Santé Publique Comité d’Ethique), Ghana (Ethical Review Committee of the Ghana Health Service and Ethical and Protocol Review Committee of the College of Health Sciences, University of Ghana), Kenya (Kenyatta National Hospital/University of Nairobi (KNH/UoN) Ethics and Research Committee), Malawi (College of Medicine Research Ethics Committee (COMREC)), Mozambique (Comité Nacional de Bioetica para e Saude, Ministerio de Saude), Nigeria (Federal Capital Territory Health Research Ethics Committee Research Ethical Review Committee, Oyo State and State Health Research Ethics Committee of Ondo State) and Uganda (Mulago Hospital Research Committee Uganda National Council for Science and Technology). All women participating in exit interviews provided informed consent.

